# Methodological proposal to explore and design future health innovation policies and opportunities for Responsible Research and Innovation (RRI)

**DOI:** 10.1101/2021.05.10.21256984

**Authors:** Arturo Argüello, Alicia Ortega-Rodriguez, Pablo Álvarez

**Affiliations:** Oficina de Transferencia de Tecnología del Sistema Sanitario Público de Andalucía. Fundación Pública Andaluza Progreso y Salud; Instituto de Investigación Biosanitaria ibs.GRANADA; Fundación para la Investigación Biosanitaria de Andalucía Oriental: FIBAO

**Keywords:** Health Innovation, Patents, Responsible Research and Innovation, Health Research Policies

## Abstract

Responsible Research and Innovation (RRI) seeks to align the research results with the needs of society to respond to current and future problems, also encompassing financial instruments, innovative public policies, and the distribution of resources. These policies should prioritize research in those areas with the greatest impact on society, and particularly in health research, this impact should be focused on responding to clinical problems of the population rather than economic impact. A methodology is proposed that uses information from open sources to identify discrepancies between the results of the most translational research of a region and its health needs, in order to provide useful information for the formulation of innovation policies.

## Introduction

Responsible Research and Innovation (RRI) was promoted by the European Commission in November 2014 through the Rome Declaration on RRI, calling on member states and their organizations and institutions to focus and prioritize RRI, declaring it essential for sustainable innovation and highlighting the responsibility of scientific achievement to be able to meet the needs and expectations of society. The statement also included the need to search for methodologies to design future innovation policies^1^.

Since the middle of the last century, patents have been used as a source of information for evaluating science and technology activities, and its use has increased as data has become more accessible electronically^2^. However, such information is imperfect (for example, not all innovations or inventions are patented), and its limitations should be considered when using it to formulate R&D&I policies, so it should preferably be combined with other indicators^3^.

In the biomedical field, the clinical application of most technologies requires substantial investment in research and development, so the use of industrial property rights, including patents, usually accompanies the development of these technologies from the earliest stages to reduce the risk of not maximizing the investment. This makes patents in this field especially useful for the identification of new technologies and technological changes, the analysis of technological trends, the analysis and monitoring of competitors, and the support for decision making in the planning and management of R&D&I^4,5^.

The information contained in clinical trials is frequently used for competitive intelligence analysis in the pharmaceutical field, together with market and patent information^6^, and it is also used for reviews. Since the promulgation of the Helsinki declaration in 1964, and also promoted by the editors of scientific journals^7^, the different countries have made the registration of information from human clinical trials mandatory by law. The WHO established a minimum number of data to be recorded^8^, which constitutes an open source of information that can be used for technological intelligence.

The International Classification of Diseases (ICD) established by the WHO^9^, allows the standardization for reporting diseases and health conditions, facilitating the comparison of mortality and morbidity statistics worldwide.

In this work we describe a methodology for obtaining information that will allow the definition of future health innovation policies, for the identification of research opportunities and prioritization of R&D&I funding through the prioritization of the projects that best suit the biomedical technologies of interest detected.

## Methods

For the analysis, we used cause-of-death and hospital morbidity per 100.000 population 2018 statistical data, classified in accordance with the International Classification of Diseases, 10th Revision (ICD–10), and extracted from the National Institute of Statistics (INE).

The patent search was performed according to the International Patent Classification (IPC), defining the appropriate classes for each ICD-10. The search was carried out in Global Patent Index (GPI) (https://www.epo.org/searching-for-patents/technical/espacenet/gpi.html), and the period analyzed was from January 1, 2009 to December 31, 2019. Patents filed by residents of Andalusia were searched in Invenes (https://www.oepm.es/es/Bases_de_Datos_Invenciones.html), using the same classification.

The clinical trial data for each therapeutic area, related to each geographical area - Andalusia, Spain and worldwide - and for the dates established in the study were obtained from the following registries: http://www.anzctr.org.au/ (Australia); http://www.chictr.org/ (China); http://www.clinicaltrials.gov (USA); https://www.clinicaltrialsregister.eu (EU); http://ctri.nic.in (India); http://www.germanctr.de (Germany); http://www.ensaiosclinicos.gov.br (Brazil); http://www.irct.ir (Iran); http://isrctn.org (United Kingdom); http://registroclinico.sld.cu (Cuba); http://www.umin.ac.jp/ctr/index/htm (Japan); http://www.trialregister.nl/trialreg/index.asp (The Netherlands).

The results obtained are shown in Table 1 and have been represented as percentages of the total analyzed in Figure 1. The data from the study can be consulted at (Reserved DOI: 10.17632/m3gtm59vfg.1).

**Table 1.** Raw data extracted from INE databases, Gobal Patent Index and clinical trial registries for each ICD-10. In parentheses, percentage with respect to the total for each column.

| ICD-10 | Andalusia<br>mortality | Andalusia<br>morbidity* | Spain<br>mortality | Spain<br>morbidity* | Andalusia<br>patents | Spain<br>patents | World<br>patents | Andalusia<br>CTs | Spain CTs | World CTs |
| --- | --- | --- | --- | --- | --- | --- | --- | --- | --- | --- |
| **Infectious** | **1176** | **208** | **6398** | **245** | **43** | **414** | **97566** | **345** | **923** | **19810** |
|  | (1,7%) | (3,5%) | (1,6%) | (3,2%) | (9,9%) | (12,2%) | (12,2%) | (10,0%) | (10,5%) | (10,5%) |
| **Tumors** | **18261** | **832** | **112714** | **991** | **82** | **584** | **107451** | **1400** | **3099** | **43114** |
|  | (27,2%) | (14,1%) | (28,2%) | (13,0%) | (18,9%) | (17,2%) | (13,4%) | (40,6%) | (35,3%) | (22,9%) |
| **Blood** | **297** | **76** | **1946** | **92** | **18** | **297** | **73034** | **125** | **362** | **6196** |
|  | (0,4%) | (1,3%) | (0,5%) | (1,2%) | (4,1%) | (8,7%) | (9,1%) | (3,6%) | (4,1%) | (3,3%) |
| **Endocrine** | **2268** | **145** | **13465** | **188** | **32** | **284** | **62163** | **299** | **697** | **19675** |
|  | (3,4%) | (2,5%) | (3,4%) | (2,5%) | (7,4%) | (8,4%) | (7,8%) | (8,7%) | (7,9%) | (10,4%) |
| **Mental disorders** | **2447** | **151** | **22376** | **253** | **11** | **120** | **15186** | **49** | **213** | **9343** |
|  | (3,6%) | (2,6%) | (5,6%) | (3,3%) | (2,5%) | (3,5%) | (1,9%) | (1,4%) | (2,4%) | (5,0<br>%) |
| **Nervous System** | **4221** | **186** | **26279** | **259** | **93** | **491** | **71864** | **338** | **966** | **29147** |
|  | (6,2%) | (3,2%) | (6,6%) | (3,4%) | (21,4%) | (14,4%) | 9,0%) | (9,8%) | (11,0%) | (15,5%) |
| **Circulatory** | **22706** | **1102** | **120859** | **1310** | **40** | **251** | **61356** | **268** | **763** | **17684** |
|  | (33,7%) | (18,7%) | (30,2%) | (17,1%) | (9,2%) | (7,4%) | (7,7%) | (7,8%) | (8,7%) | (9,4%) |
| **Respiratory** | **8375** | **888** | **53687** | **1359** | **7** | **152** | **45427** | **163** | **449** | **9618** |
|  | (12,5%) | (15,1%) | (13,4%) | (17,8%) | (1,6%) | (4,5%) | (5,7%) | (4,7%) | (5,1%) | (5,1%) |
| **Digestive** | **4028** | **1097** | **21689** | **1306** | **20** | **167** | **88448** | **183** | **516** | **13251** |
|  | (6,0%) | (18,7%) | (5,4%) | (17,1%) | (4,6%) | (4,9%) | (11,1%) | (5,3%) | (5,9%) | (7,0%) |
| **Skin** | **304** | **87** | **1826** | **119** | **56** | **300** | **72556** | **51** | **164** | **6456** |
|  | (0,4%) | (1,5%) | (0,5%) | (1,6%) | (12,9%) | (8,8%) | (9,1%) | (1,5%) | (1,9%) | (3,4%) |
| **Osteomuscular** | **882** | **536** | **5205** | **768** | **19** | **206** | **49943** | **148** | **392** | **8001** |
|  | (1,3%) | (9,1%) | (1,3%) | (10,1%) | (4,4%) | (6,1%) | (6,2%) | (4,3%) | (4,5%) | (4,2%) |
| **Genitourinary** | **2360** | **574** | **13941** | **750** | **13** | **134** | **54148** | **83** | **231** | **6175** |
|  | (3,5%) | (9,8%) | (3,5%) | (9,8%) | (3,0%) | (3,9%) | (6,8%) | (2,4%) | (2,6%) | (3,3%) |

\* Hospital Morbidity Rates per 100.000 inhabitants.

**Figure 1.**
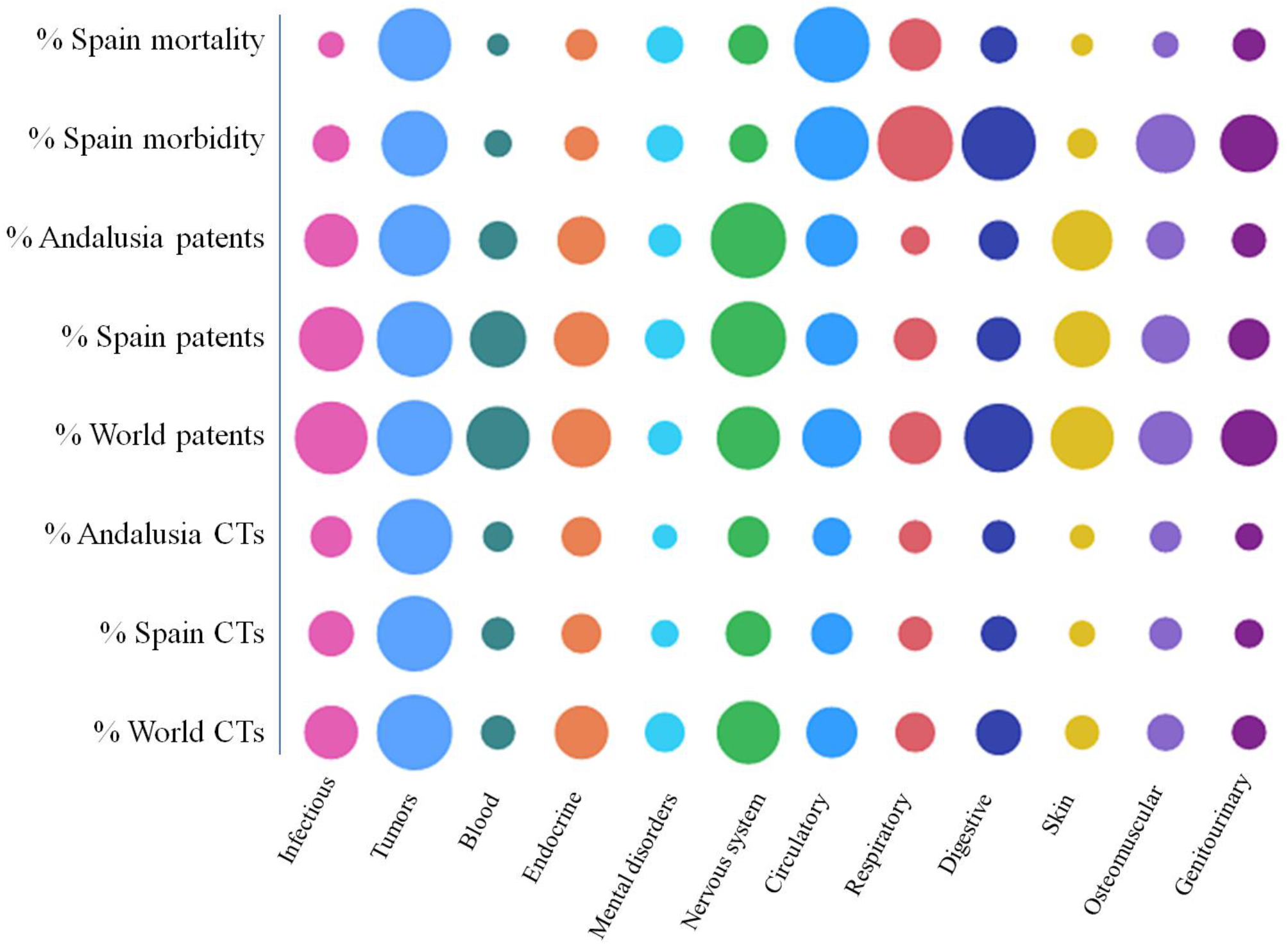
Graphical representation of the percentages of each variable proportional to the area of each circle for each ICD-10.

## Results

Figure 1 shows that circulatory diseases are the main cause of mortality and morbidity in Spain; however, the number of patents and clinical trials, indicators of investment in this area, is not equivalent. In neoplasms, the second leading cause of death, a correlation is observed. In diseases of the digestive system, genitourinary system and respiratory system, there is a deviation between morbidity and the clinical trials registered. With respect to skin, endocrine and nervous system diseases, a greater inversion is observed, that is not in accordance with the mortality and morbidity data.

## Discussion

The same methodology could be applied at a more specific level, for example, to determine the R&D&I situation in specific diseases that are particularly prevalent in a region.

Specific developments and mechanisms have been generated to include in the research and innovation process the social utility of science and its potential impacts, enabling greater assurance of translational research. Currently, RRI is a topic that involves researchers, companies, scientific institutions, and research funding agencies. It is therefore proposed to use the term collective responsibility^10^ to define this concept and to implement the positive impacts of research and innovation.

The collection, correlation and processing of publicly accessible data such as patents and clinical trials can be used to obtain useful information^11,12^, particularly in the formulation of regional innovation policies^13,14^. By adding ICD statistics, it is possible to identify technological fields where, for reasons of public health, it may be interesting to prioritize research funding from the public sector, so that public research and innovation have an impact on clinical care, fostering a culture of RRI in the scientific community and facilitating the transfer of scientific achievements to society.

## Data Availability

The data that support the findings of this study are openly available in Mendeley at DOI: 10.17632/m3gtm59vfg.1.

http://www.doi.org/10.17632/m3gtm59vfg.1

## Authorship contributions

A. Argüello, A. Ortega-Rodríguez and P. Álvarez conceived the study. A. Ortega-Rodríguez carried out the searches in the databases. All authors contributed ideas, reviewed the drafts, and approved the final version.

## Financing

This work has been carried out with the help AT-6052, entitled “*Prospección de Campos Tecnológicos en Biomedicina mediante el Análisis Estratégico de Patentes*”, funded by the Consejería de Economía, Conocimiento, Empresas y Universidad and co-financed by the European Regional Development Fund (ERDF).

## Conflicts of interest

None.

## Notes

### Competing Interest Statement

The authors have declared no competing interest.

### Funding Statement

This work has been carried out with the help AT-6052, entitled "Prospeccion de Campos Tecnologicos en Biomedicina mediante el Analisis Estrategico de Patentes", funded by the Consejeria de Economia,
Conocimiento, Empresas y Universidad and co-financed by the European Regional Development Fund (
ERDF)

### Author Declarations

Ethics approval was not required for this study

